# *“If I am free from diabetes, that itself will be the happiest thing”*: A convergent mixed methods study of the lived experiences of young adults with type 2 diabetes in Mysore district, India

**DOI:** 10.1101/2024.02.23.24303271

**Authors:** Nikhita R. Gopisetty, Kiranmayee Muralidhar, Nagalambika Ningaiah, Rani Chinnappa, Mia Buono, Poornima Jaykrishna, Purnima Madhivanan, Sumedha G. Ariely, Eve S. Puffer

**Author notes:** Corresponding author (NRG). These authors contributed equally to this work (Sumedha G. Ariely, Eve S. Puffer).

## Abstract

Type 2 diabetes (T2D) has been occurring at younger ages of onset around the world. India’s population accounts for nearly 20% of the global disease burden. This study investigated the occurrence of depressive symptoms and qualitatively explored the lived experiences of 20 young adults living with T2D under the age of 35. We conducted a convergent mixed-methods study with the Patient Health Questionnaire (PHQ-9) and semi-structured interviews from June 2022 to July 2022 in Mysore district, India. Guided by the World Health Organization’s Commission on Social Determinants of Health conceptual framework and biopsychosocial frameworks, areas of inquiry included knowledge and perception about T2D, accessibility of healthcare resources, T2D-related self-care activities, and the impact of the condition on their daily life. Interviews were debriefed by the research team and analyzed thematically using NVivo 12. Participants were aged between 21 and 35 (mean: 30.8, SD: 4.2) and the majority were female (75%). Overall, 55% reported mild depression symptoms, 15% reported moderate to moderately severe depression symptoms; 5 participants (25%) reported suicidality. Sex, living in rural Mysore district, socioeconomic status, T2D duration, family history of T2D, T2D-induced complications, and T2D-related self-care behaviors were associated with depressive symptoms. Thematic analysis revealed 1) low knowledge about T2D, 2) substantial interpersonal and internalized stigma for having T2D at a young age, 3) financial and time constraints to seek and receive care, 4) self-perception as burdens to family members due to the cost and stress of living with T2D, 5) competing priorities with work and family, and 6) the power of social support in managing T2D. These themes were consistent across the sample, regardless of severity of depressive symptoms. Awareness campaigns and peer support programs may help reduce depressive symptoms and increase self-efficacy in this population.

## Introduction

Diabetes is a global epidemic – in 2019, over 450 million adults were estimated to be living with type 2 diabetes (T2D) and this number is predicted to exceed 700 million by 2045 [1]. Almost 80% of people living with T2D live in low- and middle-income countries (LMICs); over 77 million people with T2D live in India, of which over 50% are undiagnosed [2]. Diabetes is occurring at a younger age of onset and lower BMI levels across India and around the world. In rural Pondicherry, India, one third of new diabetes cases occur in people below 40 years old [3]. In younger populations living with T2D, comorbidities also occur at a younger age, including fatty liver disease and systolic hypertension [4]. Diabetes-related complications impact the physical, mental, and social well-being of patients, placing a burden on the healthcare system [5]. Sub-Saharan Africa, South America, and parts of Asia have yet to experience the epidemiological transition from communicable diseases to non-communicable diseases – India is an example to learn from before this occurs [6].

Prior research has established a bidirectional relationship between diabetes and depression [7–9]. In urban-based clinics around India, researchers found that one-quarter to one-third of patients with T2D were depressed. They predict that the prevalence of depression in people living with T2D might be higher in LMICs than high income countries [10]. Depressive disorders in people living with T2D are associated with the female gender, young age of onset, lower education levels, worse glycemic control, a lack of physician advice about lifestyle modifications, and low social support [7,8,11–13]. Lower-income populations exhibit higher rates of depression, due to a lack of access to healthcare, higher likelihood of developing complications, and a low social awareness [14]. The comorbidity of depression and chronic disease is associated with worse physical disorder outcomes and quality of life [7,13].

Previous research has shown younger age to be significantly associated with diabetes distress in South India [15]. Young adults living with diabetes have been shown to have lower self-esteem than those without, due to social perceptions and preconceived notions about the condition [16]. Key emotions in young diabetics discussing their disease included denial, anger, depression, and guilt. They feared people would treat them differently and perceive them as sick, and experienced unsolicited advice and instructions about household remedies from family members and peers [17]. Diabetes occurring at a younger age is also associated with significant social stigma, due to societal and cultural misconceptions about the disease [18].

As T2D becomes more prevalent in younger populations, it is crucial to understand the various barriers to diabetes management and quality of life that they face. A conceptual model, derived from the WHO’s Commission on Social Determinants of Health (CSDH) conceptual framework [19] and the biopsychosocial model [20], was developed to guide this study (Fig 1). The CSDH framework presents the relationships between structural and intermediary social determinants of health and equity in health and well-being [19]. The biopsychosocial model of health and illness allows researchers to model any disease caused and impacted by biological, psychological, and social factors, revealing interactions between the mind and body [20]. The conceptual model below maps out various pathways between sociodemographic characteristics, structural and intermediary determinants, T2D-related self-care behaviors, physical health outcomes, and quality of life. Intermediary determinants -- including health literacy, access, social factors, and psychological factors -- are influenced by an individual’s environment and directly impact self-care behaviors and health outcomes. The purpose of this study is to apply this conceptual model to explore the impact of intermediary determinants of health on the prevalence of depressive symptoms and lived experiences in young adults living with T2D in Mysore district, Karnataka, India.

**Fig 1.**
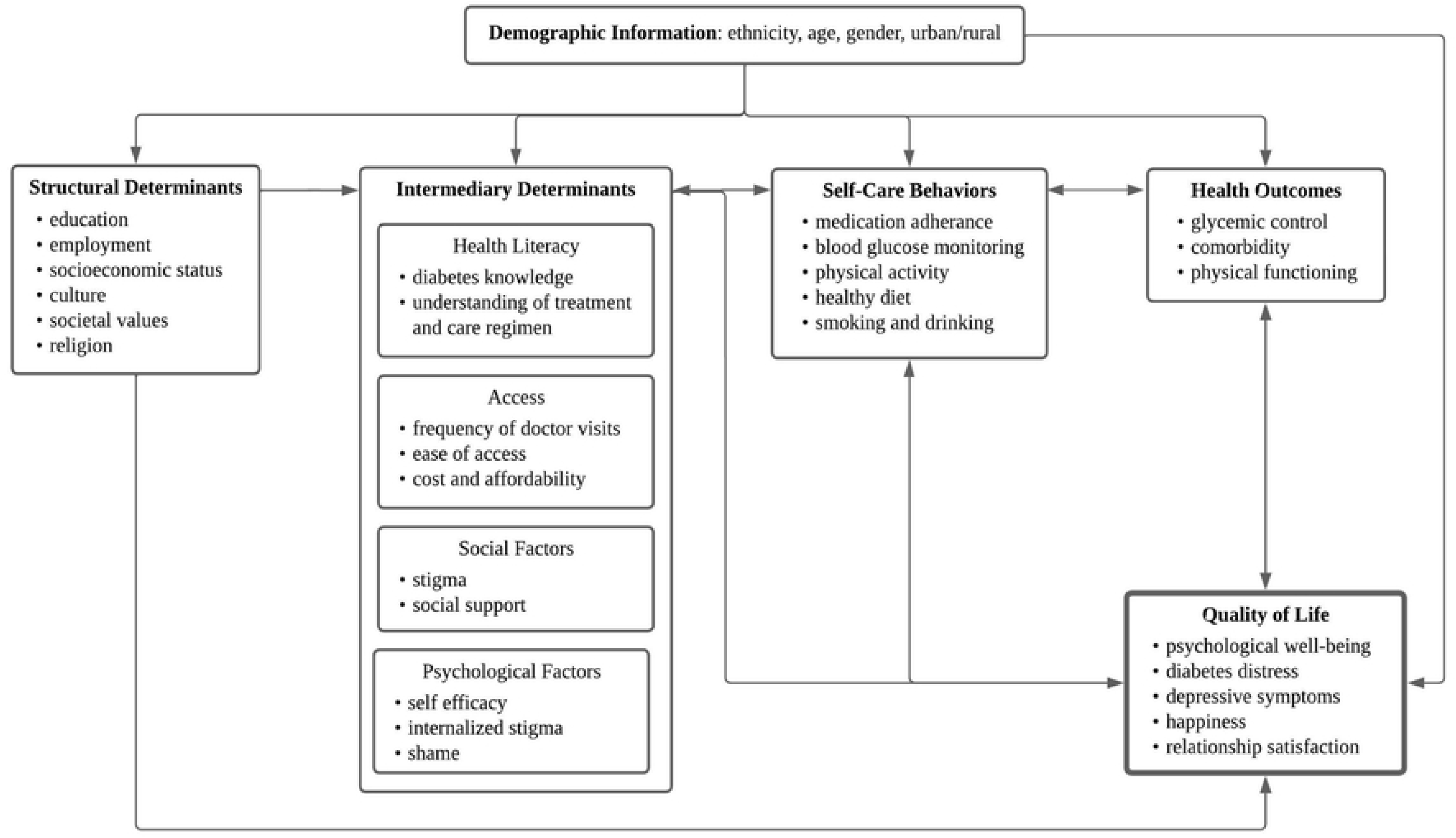
Conceptual model.

## Methods

### Study design

We used a convergent mixed-methods approach, with an emphasis on deeper understanding and analysis of qualitative themes [21]. Qualitative and quantitative data were collected concurrently and findings were interpreted with the intent of validating and describing relationships among the data. Approval was obtained from Duke Campus Institutional Review Board (2022-0358) and the Public Health Research Institute of India’s Institution Ethics Review Board (2022-01-29-64).

### Setting

The study was conducted in Mysore, a district in southern Karnataka in India between June and July 2022. The Indian National Census in 2011 reported a total population of 3,001,127 in Mysore district, with approximately 58% living in rural villages [22]. The National Family Health Survey-5 (NFHS-5) from 2019 to 2021 reported that India’s population is young, which is typical of developing countries with a low life expectancy; over 50% of India’s population is below the age of 30 [23]. The study was implemented in collaboration with the Public Health Research Institute of India (PHRII), a research and service-based organization in Mysore, Karnataka. PHRII has been conducting community-based research, providing health services, and training public health professionals in Mysore district for the past 15 years.

### Author positionality

It is important to recognize the authors’ positionality and, therefore, their interpretation of the data. The research team consisted of public health practitioners, academic researchers, physicians, administrative staff, a graduate student, and an undergraduate student. Seven members of the research team come from Indian backgrounds, two born in the United States and five born in India. Two members of the research team are United States-born White Americans with experience designing and implementing research projects in global settings. All authors worked as a team to ensure the study was guided by their collective cultural knowledge and expertise. Positionality challenges related to sex, gender, race/ethnicity, power, socioeconomic status, and privilege were intentionally addressed throughout the research design and data collection processes through team-based reflection on how these issues might impact the interview process and data analysis.

### Eligibility and participant recruitment

Young adults with T2D were recruited using convenience sampling between June 6, 2022 and July 8, 2022. Participants were eligible to take part in the study if they were between 18 to 35 years old, self-reported to have T2D, live in Mysore district, able to speak English or Kannada, and able to provide informed consent. The research team consulted with Primary Health Centres, Accredited Social Health Activists (ASHAs), Urban Social Health Activists (USHAs), and the Karnataka Health Promotion Trust to identify potential participants who met the inclusion criteria. Alongside the ASHAs and USHAs, government-supported community health workers, the research team physically approached potential participants in their communities to inform them of the study. Written informed consent was obtained from the study participants after explaining the purpose of the study and reading through the content of the study information sheet and consent form. Of 21 eligible candidates that were approached, one refused to participate in the study. This resulted in 20 eligible participants, 10 from urban and 10 from rural Mysore district.

## Measures

### Quantitative

Participants completed a demographic questionnaire that included sociodemographic characteristics, medical history information, and details regarding their diagnosis with T2D and related management. Participants were screened for depressive symptoms using the Patient Health Questionnaire-9 (PHQ-9) [24], a brief self-report instrument of depression severity that has been translated and validated for Indian populations in various languages [25]. The screening tool uses 9 questions scored on a 4-point Likert-scale ranging from 0 (not at all) to 3 (nearly every day) to examine symptoms over the past two-week period. Individual’s PHQ-9 score was categorized: 1-4 minimal depression, 5-9 mild depression, 10-14 moderate depression, 15-19 moderately severe depression, and 20-27 severe depression [24]. A single interviewer (NN) and observing notetaker (RC) verbally administered the surveys in *Kannada* and recorded responses through an offline Qualtrics survey.

### Qualitative

A semi-structured interview guide (S1 File) was developed based on the conceptual model. This guide covered the following domains and themes: general knowledge and perception about T2D, accessibility to healthcare resources, self-care behaviors related to T2D, impact of T2D on the individual, and impact of T2D on others around them. The interview guide was developed in English by the authors (NRG, KM, PJ, SA, ESP) and subsequently translated into *Kannada* by the interviewer. The guide was back-translated into English to ensure language consistency and validity of the questions. To assess the flow and colloquially adapt the language of the interview guide, three mock interviews were conducted by the interviewer and staff members at PHRII, who understood the local language and context where the study was conducted. During each mock interview, staff members provided feedback on the language of the questions and cultural appropriateness of the topics. Subsequent changes were incorporated following each mock interview to make the wording of questions more understandable and improve the flow of the conversation.

Interviews were completed either in a quiet, private room at PHRII or the participant’s home. Interviews were conducted by a single interviewer (NN) who was trained in qualitative research methods and had experience conducting semi-structured, in-depth interviews. Interviews lasted for approximately 60 minutes and were audio recorded on a digital voice recorder. During the interviews, the interviewer used probes to encourage participants to share anything relevant to the topics discussed, beyond those laid out in the interview guide. The notetaker (RC) observed and documented key observations from each interview.

## Analysis

### Quantitative

All quantitative analyses and data visualizations were conducted in Microsoft Excel (2021) and RStudio (version 4.2.1; R Core Team 2021). Socioeconomic status was classified by the modified BG Prasad scale, using annual household income and family size to calculate per-capita income [26]. Individual’s depressive symptoms were scored by summing the total PHQ-9 values for all 9 questions, with higher scores corresponding to increased symptomatology [24].

### Qualitative

The research team debriefed each interview and discussed field notes within 48 hours of completion. Interviews were transcribed verbatim and translated to English. The transcribed interviews were reviewed by the first author (NRG) to ensure transcriptions were accurate and complete. Debrief notes were correlated to the respective transcripts upon completion. Two researchers (NRG, KM) familiarized themselves with the data and generated a codebook for thematic qualitative analysis, based on the preconceived conceptual model (Fig 1). Two researchers (NRG, MB) independently coded the transcripts and debrief notes with NVivo 12 software. Interrater reliability was established with a kappa coefficient of 0.86, indicating almost complete agreement [27]. Discrepancies in coding between the investigators were resolved through consensus with a third researcher (KM). Emergent themes and relevant sub-themes were then discussed among all authors to promote consistency and minimize bias. Thematic saturation was reached after analyzing data from the interviews, ensuring comprehensive coverage of the study’s qualitative findings.

## Results

Twenty participants with T2D participated in the study, of which 15 were female. Twelve participants were diagnosed with T2D within the past two years. The age of participants ranged from 21 to 35 years with a mean age of 30.8 (SD ± 4.2) All spoke Kannada as their first language. Seven were lower-middle class and six were middle class, according to the modified BG Prasad scale. Table 1 presents key health and demographic characteristics of the study participants, stratified by rural/urban distinction.

**Table 1.**
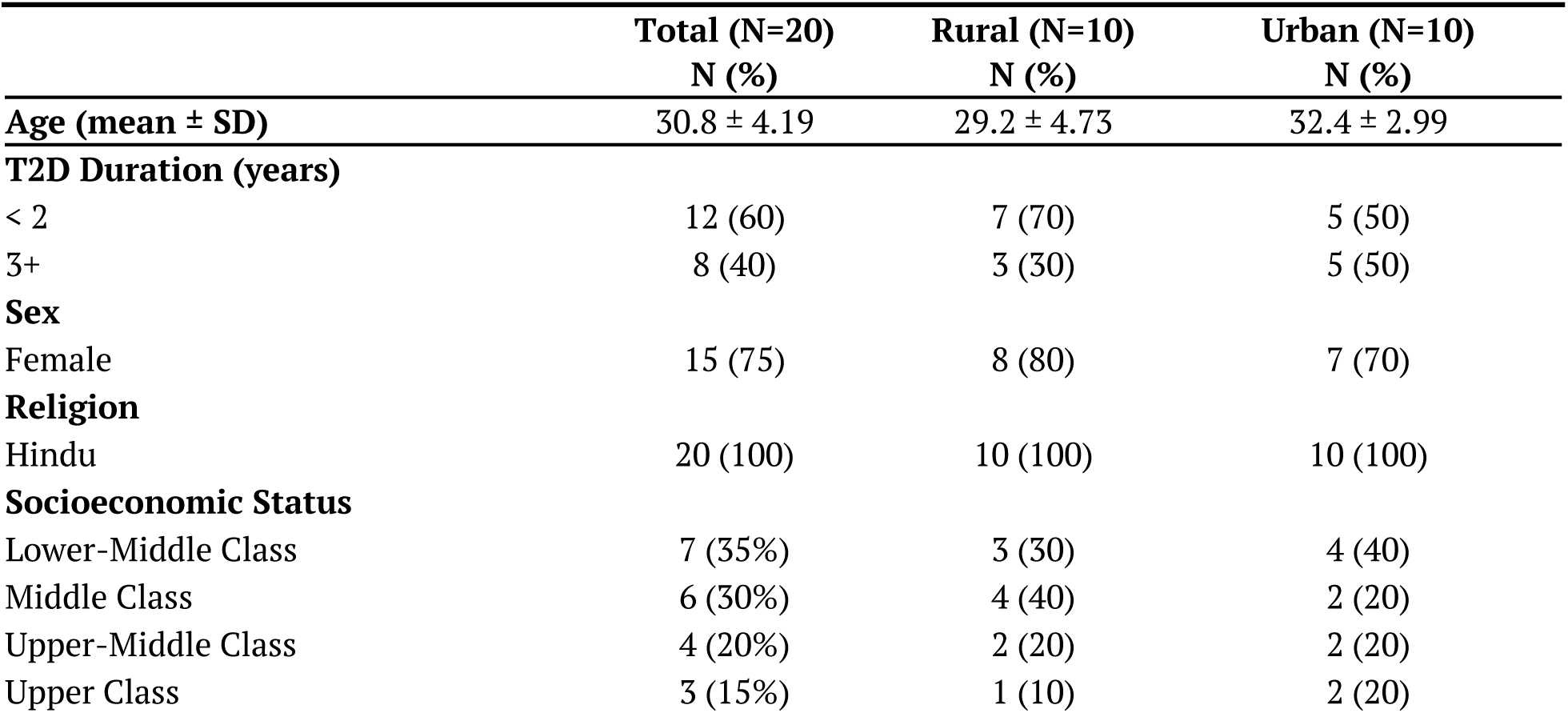
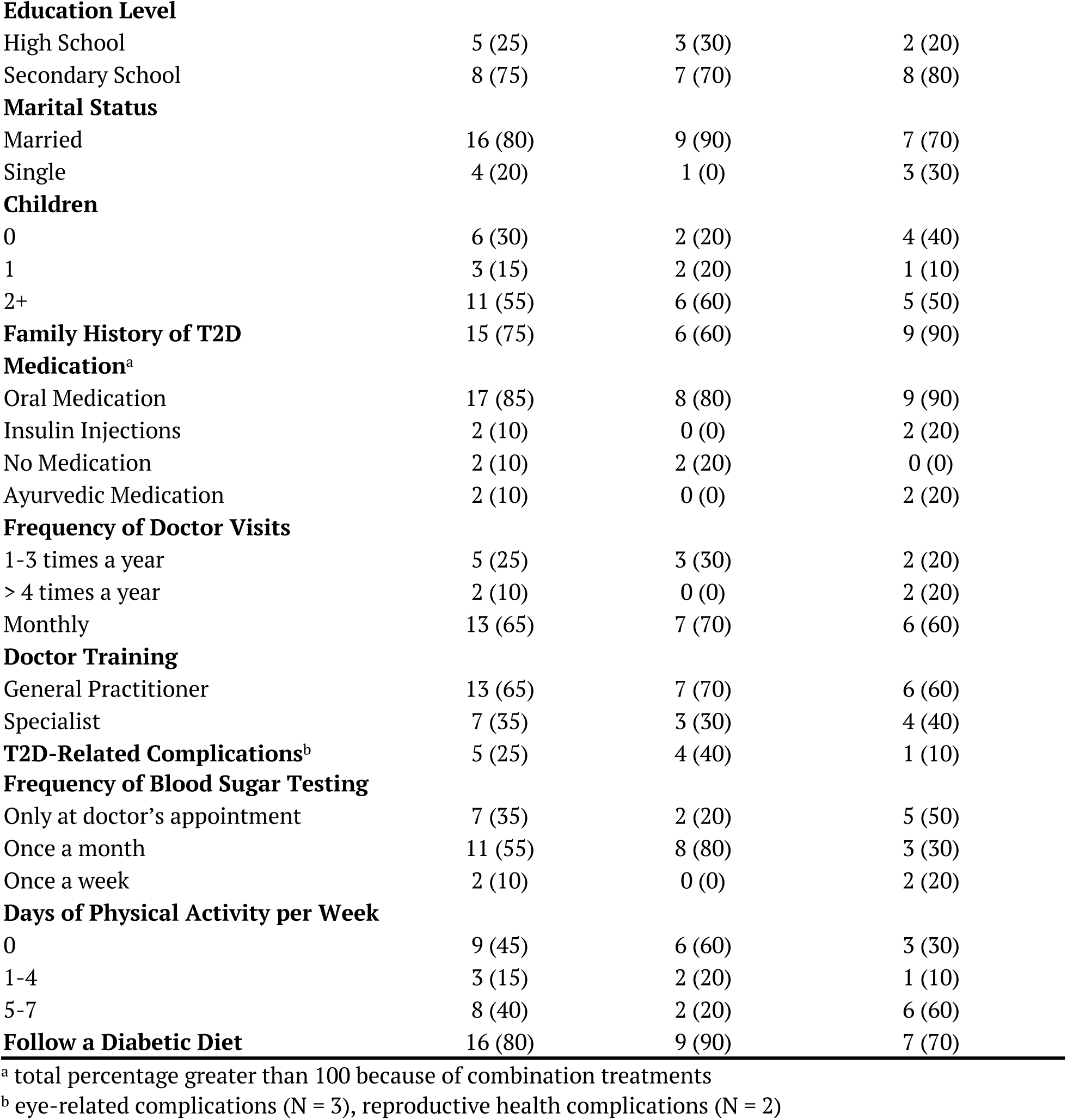
Participant sociodemographic and health behavioral characteristics.

|  | Total (N=20)<br>N (%) | Rural (N=10)<br>N (%) | Urban (N=10)<br>N (%) |
| --- | --- | --- | --- |
| **Age (mean ± SD)** | 30.8 ± 4.19 | 29.2 ± 4.73 | 32.4 ± 2.99 |
| **T2D Duration (years)** |  |  |  |
| < 2 | 12 (60) | 7 (70) | 5 (50) |
| 3+ | 8 (40) | 3 (30) | 5 (50) |
| **Sex** |  |  |  |
| Female | 15 (75) | 8 (80) | 7 (70) |
| **Religion** |  |  |  |
| Hindu | 20 (100) | 10 (100) | 10 (100) |
| **Socioeconomic Status** |  |  |  |
| Lower-Middle Class | 7 (35%) | 3 (30) | 4 (40) |
| Middle Class | 6 (30%) | 4 (40) | 2 (20) |
| Upper-Middle Class | 4 (20%) | 2 (20) | 2 (20) |
| Upper Class | 3 (15%) | 1 (10) | 2 (20) |
| **Education Level** |  |  |  |
| High School | 5 (25) | 3 (30) | 2 (20) |
| Secondary School | 8 (75) | 7 (70) | 8 (80) |
| **Marital Status** |  |  |  |
| Married | 16 (80) | 9 (90) | 7 (70) |
| Single | 4 (20) | 1 (0) | 3 (30) |
| **Children** |  |  |  |
| 0 | 6 (30) | 2 (20) | 4 (40) |
| 1 | 3 (15) | 2 (20) | 1 (10) |
| 2+ | 11 (55) | 6 (60) | 5 (50) |
| **Family History of T2D** |  |  |  |
|  | 15 (75) | 6 (60) | 9 (90) |
| **Medication<sup>a</sup>** |  |  |  |
| Oral Medication | 17 (85) | 8 (80) | 9 (90) |
| Insulin Injections | 2 (10) | 0 (0) | 2 (20) |
| No Medication | 2 (10) | 2 (20) | 0 (0) |
| Ayurvedic Medication | 2 (10) | 0 (0) | 2 (20) |
| **Frequency of Doctor Visits** |  |  |  |
| 1-3 times a year | 5 (25) | 3 (30) | 2 (20) |
| > 4 times a year | 2 (10) | 0 (0) | 2 (20) |
| Monthly | 13 (65) | 7 (70) | 6 (60) |
| **Doctor Training** |  |  |  |
| General Practitioner | 13 (65) | 7 (70) | 6 (60) |
| Specialist | 7 (35) | 3 (30) | 4 (40) |
| **T2D-Related Complications<sup>b</sup>** |  |  |  |
|  | 5 (25) | 4 (40) | 1 (10) |
| **Frequency of Blood Sugar Testing** |  |  |  |
| Only at doctor's appointment | 7 (35) | 2 (20) | 5 (50) |
| Once a month | 11 (55) | 8 (80) | 3 (30) |
| Once a week | 2 (10) | 0 (0) | 2 (20) |
| **Days of Physical Activity per Week** |  |  |  |
| 0 | 9 (45) | 6 (60) | 3 (30) |
| 1-4 | 3 (15) | 2 (20) | 1 (10) |
| 5-7 | 8 (40) | 2 (20) | 6 (60) |
| **Follow a Diabetic Diet** |  |  |  |
|  | 16 (80) | 9 (90) | 7 (70) |

<sup>a</sup> total percentage greater than 100 because of combination treatments
<sup>b</sup> eye-related complications (N = 3), reproductive health complications (N = 2)

### Depressive symptoms

Table 2 presents the results of the PHQ-9 screening for the participants. While a small sample size, rates of symptoms were high. Of the 20 participants, over half (55%, N = 11) of the study participants had scores in the mild depression range, 15% (N = 3) in the moderate to moderately severe depression range. Five participants reported having suicidal thoughts in the past 2 weeks -- 3 with scores in the mild depression range and 2 in the moderately severe depression range. The highest scores were associated with the following 3 prompts: 1) Little interest or pleasure in doing things; 2) Feeling down, depressed, or hopeless; and 3) Feeling tired or having little energy.

**Table 2.**
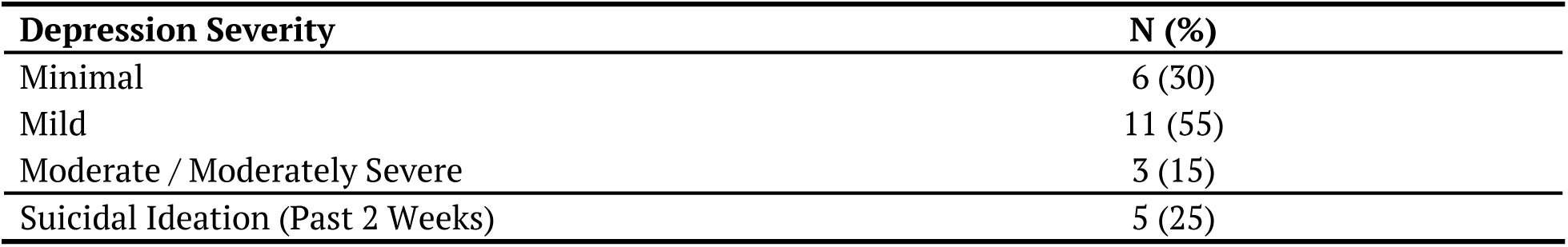
Depression severity (PHQ-9)

Fig 2 visually presents the relationships between covariates and depressive symptoms. Participants who identified as female, resided in rural Mysore district, and belonged to a lower socioeconomic status were more likely to have depressive symptoms. Participants who were diagnosed with T2D within 2 years, did not have a family history of T2D, faced T2D-related complications, adhered to a diabetic diet, and neglected physical activity in their routine were also more prone to depressive symptoms.

**Fig 2.**
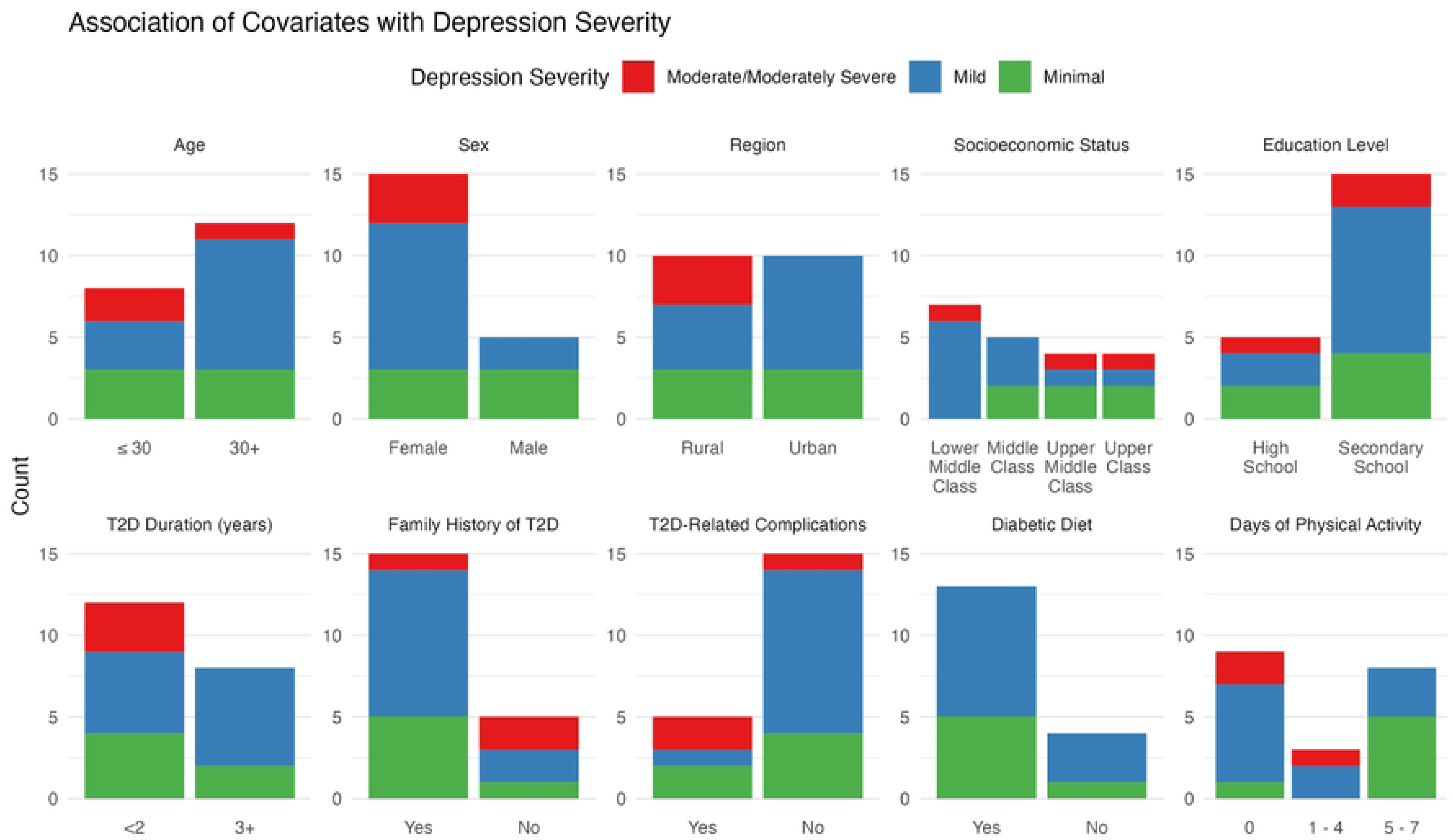
Association of covariates with depressive symptoms.

## Intermediary determinants of diabetes self-management

### Health literacy

When asked how many people in their community have diabetes, participants predicted an average of 70% (SD 16.8%), indicating the seemingly high prevalence. In 2021, the prevalence of diabetes in India was reported at 9.6%, drastically different from the participants’ perception [2]. Participants across rural and urban Mysore district showed varied levels of awareness about the prevalence of diabetes in younger populations. In rural Mysore district, 90% of participants stated that they did not know other people their age living with the condition:

> “*People with very young age will not be affected by diabetes… I think I am exceptional.*” (P10, Female, PHQ-9 = 6, Mild Depression)

However, in urban Mysore district, participants reported greater awareness about the rising prevalence of diabetes at younger ages:

> *“Now almost everyone is becoming diabetic at a younger age. If the parents have it, the kids might acquire it too, and with that, the food we consume is also not good. So, from that as well, people are getting sugar [diabetes] soon.”* (P15, Female, PHQ-9 = 6, Mild Depression)

Overall, participants reported having a lack of trustworthy sources to learn about diabetes. As a result, 7 (35%) felt scared to ask questions to better understand their biological condition, due to false information or societal stigma:

> *“Few of them will say that food may also cause diabetes, but few say that it is not because of food. I don’t know whom I should listen and follow.”* (P10, Female, PHQ-9 = 6, Mild Depression)

Participants reported that their healthcare providers recommended them to maintain a diabetic diet, perform physical activity regularly, and to avoid stress to manage their condition. However, due to the large quantity of patients in the clinic, participants rarely experienced individualized care. One participant stated:

> *“My doctor won’t specifically mention everything, they will just say to have proper food intake and be calm without worrying about anything. Sometimes I feel bad that they will not tell me properly about diabetes.”* (P9, Female, PHQ-9 = 15, Moderately Severe Depression)

All of the participants knew that diabetes-related complications exist, but the majority (65%) were not adequately informed about the complications and ways to prevent them:

> *“They [doctors] said that if there is a wound in the leg, they will amputate it and there will be loss of vision. If there is a wound in the hand, they will amputate it.”* (P11, Female, PHQ-9 = 9, Mild Depression)

Nearly all participants (90%) explicitly stated that they do not want to take medication for the rest of their lives. These perceptions are influenced by participants’ community members and even healthcare professionals.

> *“They have said to control diabetes by taking proper food and stopping the tablets. Even our family doctor suggested I should stop taking tablets and not develop a habit of taking them forever.”* (P13, Male, PHQ-9 = 0, Minimal Depression)

Twelve participants (60%) used language about “addiction” when referring to their diabetes medication, claiming that they do not want to become addicted to the tablets:

> *“If I get addicted to tablets, it will be difficult. I want to control it [diabetes] in food. So, I am not taking any tablets.”* (P11, Female, PHQ-9 = 9, Mild Depression)

### Access

In rural Mysore district, participants reported constant crowds at the government hospitals, requiring patients to enter a queue just to meet with the doctor. This comes alongside various challenges – women must consider who will accompany them to their appointments, working professionals must request leave from their jobs in advance, and all must find transportation to and from the clinic. One participant described her challenges with seeing a healthcare provider:

> *“Sometimes doctors will not be there. One day I waited for so many hours but the doctor did not come to the hospital. So I came back home, if he is not there I will go the next day.”* (P10, Female, PHQ-9 = 6, Mild Depression)

Seven participants (35%) stated that they prefer getting their healthcare at private clinics because they believe that if they are paying for it, the service must be better: *“I feel that whatever the money I spend, my health should be fine and good. So I will be happy.”* (P2, Female, PHQ-9 = 15, Moderately Severe Depression).

With a greater presence of healthcare facilities, there are more opportunities to meet with a doctor in urban Mysore district. Some doctors in the urban region even adjust their hours to be available after the workday. However, participants reported that crowds are still present at the clinics. Due to the crowds and long queues, some patients reported feeling like a burden if they take too much time with the doctor, which interferes with their own care. Some participants described:

> *“There will be so many patients like me, if I take too much time to discuss my problems, it will get very late for other patients. I will be troubling them. So I will think that I can talk to him the next time when I go for a checkup.”* (P12, Female, PHQ-9 = 7, Mild Depression)

Blame from healthcare providers emerged as a theme, illustrating some of the complicated and diverse power dynamics between patients and their providers. Over half of participants in urban Mysore district reported that they experienced blame from their providers if their diabetes was not under control.

> *“She [doctor] actually scolds me to take proper care of my health. She also mentioned that due to my negligence I am unable to take proper care of my health. If I don’t start taking care of it she has warned me to not visit the clinic.”* (P20, Female, PHQ-9 = 7, Mild Depression)

Despite these challenges, some patients are willing to invest the time to take care of their health. They navigate the difficulty with positive attitudes:

> *“If we want to get the medicines we should wait, it’s not like they should only treat me, there will be so many people like me. Even they will have some problem, we can’t say that our time is getting wasted and blame doctors. If we are unable to contact him today, we can visit the next day.”* (P6, Male, PHQ-9 = 0, Minimal Depression)

#### Financial burden

Due to competing financial priorities, most participants (65%) are unable to afford their diabetes medication. All of the participants with children mentioned that they did not want their diabetes to interfere with their children’s education:

> *“I cannot spend my wages on my tablets. I have to take care of my children’s education.”* (P18, Female, PHQ-9 = 6, Mild Depression).

Some participants reported skipping doses if they do not have the means to purchase it, and others choose to not spend their limited expendable income on their medications. One participant reported that she preferred government-subsidized medications, but they are not available in her area. Three participants reported that they try to maximize their limited supplies of medication by using it as a reactive mechanism, rather than a preventative mechanism. They explained that they wait to take the medication until they experience symptoms of high blood sugar levels:

> *“When I get the burning sensation in my feet, I come to know that my sugar level is high. Then, I take one tablet and gulp it. That is all.”* (P11, Female, PHQ-9 = 9, Mild Depression)

Under financial constraints, people living with diabetes find themselves having to choose between speaking to their doctor or purchasing their medication due to the costs involved. In these scenarios, they are managing their condition blindly, until they can visit their doctor again.

> *“I will not go for a blood test every month because of financial problems. I will visit the private hospital so the cost will be more, so I will just purchase tablets.”* (P9, Female, PHQ-9 = 15, Moderately Severe Depression)

For some, their diabetes is still poorly controlled, so they questioned the purpose of spending their money:

> *“I purchase the tablets and I give the amount for appointments. Even though I spend so much, my [blood] sugar will not get reduced. I am fed up with it.”* (P5, Female, PHQ-9 = 13, Moderate Depression)

### Social factors

During the interviews, all of the participants described encounters with people who believed that diabetes was communicable, both in the rural and urban regions. This misconception was so well-established in certain communities that 5 (25%) participants reported never seeing their neighbors again after they shared their diagnosis.

> *“Once I was going out to the hospital and my neighbor asked what the problem was. I said I have diabetes. They thought it was contagious because I have got diabetes at an early age. They decided not to have any contact with us after that.”* (P8, Female, PHQ-9 = 9, Mild Depression)

All of the participants reported experiences of teasing from community members for having diabetes at a young age. Despite its ubiquity, it is still a taboo topic. The young adults described feeling embarrassed to share their diagnosis because others talk and laugh about them, even if they “*pretend to act like they care*” (P1, Female, PHQ-9 = 9, Mild Depression):

> *“I do feel embarrassed to share. People who stay in the village will tease me, saying that at an early age I have got diabetes. Some will understand, but not all will get to know our problems.”* (P4, Female, PHQ-9 = 1, Minimal Depression)

Fear of this community gossip and stigma often deterred participants from going to public health camps or visiting the community health workers, out of fear that someone would “*watch and talk behind [her] back to the neighbors*” (P10, Female, PHQ-9 = 6, Mild Depression).

> *“The ASHA (Accredited Social Health Activist) workers ask me to get the sugar tested in camps, but I do not go and I do not like to go. When I have to go for a checkup, I just tell others that I am going out. I will never disclose about sugar [diabetes] checkup because I do not like to say it. The way they look at us is different. So, I do not wish to disclose. That is sad.”* (P18, Female, PHQ-9 = 6, Mild Depression)

Due to the prevalent social stigma, the 3 unmarried participants described their fear of not finding a partner because of their diagnosis with diabetes. One married participant shared that she did not inform her partner of her diagnosis until after the wedding ceremony was complete, out of fear that his family would no longer approve of the union.

#### Barriers to diabetes self-management

Sixteen participants (80%) self-reported following a diabetic diet, different from the rest of their family members, increasing the burden on whoever is cooking the food. Many participants spoke about how they would rather eat the same food as everyone else than burden others. Some discussed the concept of portion control, while others just eat the same to fit in.

> *“When we are all together, I can’t ask for separate food. I should have what all others are having.”* (P3, Female, PHQ-9 = 0, Minimal Depression)

Two participants (10%) described forgoing their recommended diabetic diet due to fear of what others would think about them. If they were to change their lunches at their workplace, they were afraid that their condition would be revealed to their employer and coworkers:

> *“I will have breakfast early in the morning. But for my workplace, I will carry rice. They will think I am someone special if I don’t, so I have to carry rice for lunch.”* (P3, Female, PHQ-9 = 0, Minimal Depression)

Some female participants do not feel comfortable going for a walk or performing physical activity in public by themselves, especially at night. Then, they are reliant on their husbands to accompany them on their walks.

> *“In the evening it will be late and it becomes dark, so I will not go alone.”* (P1, Female, PHQ-9 = 9, Mild Depression)

#### Importance of positive social support

The presence of familial support is known to be an important factor in diabetes management and quality of life. Some female participants reported encouraging support from their husbands to visit the hospital and reduce stress:

> *“My husband will say that no matter how much money or time we spend, I should be healthy without any problems. He is even ready to take me to the other hospital. He is not worried about money.”* (P10, Female, PHQ-9 = 6, Mild Depression)

One participant described her close relationship with a friend who also has T2D. Her friend was successfully managing her glycemic control and served as an inspiration and motivation for her to take care of her own health.

> “*One of my friends has got diagnosed with diabetes. She is having regular spouts in her daily food routine and she is doing some exercise, going for a walk to maintain her health. I feel if I do the same I may also have a proper control in sugar level and can be healthy.*” (P10, Female, PHQ-9 = 6, Mild Depression)

### Psychological factors

Feelings of sadness and depressive symptoms after diagnosis with diabetes was common amongst study participants. Most participants (75%) described a sentiment of “why me?” and “what else could happen to me?”

> *“I felt sad and worried about the occurrence of diabetes at this young age, and also felt sad that I cannot eat most of the things which I like.”* (P1, Female, PHQ-9 = 9, Mild Depression)

Some participants relate their overall happiness to their diabetes diagnosis and glycemic control, resulting in a lack of individual control over their general well-being.

> *“If the sugar level is high, I will be very tensed. If it’s normal, I will be happy and calm.”* (P10, Female, PHQ-9 = 6, Mild Depression)

A feeling of worry due to living with diabetes at a young age was common across the study participants. This ranges from impeding routine activities, not being able to manage their glycemic control, and troubling others in their family.

> *“I got it from my mom, and I don’t want it to happen to my kids because of me. I have two daughters. What if they have problems when they get married and have complications during their pregnancies? Now, people are getting diabetic during their pregnancy. I had two kids and after that I got diabetes, but people in the present generation are getting diabetes during pregnancy. I am afraid about that.”* (P15, Female, PHQ-9 = 6, Mild Depression)

All but one participant expressed fear about future complications and early death due to poorly controlled diabetes:

> *“I feel that I may die early, I will be worried that there will be no one to take care of my daughter. They also say that there will be more consequences like kidney stone, heart disease, and also some may lose their eyesight. So I am worried about all of these.”* (P8, Female, PHQ-9 = 9, Mild Depression)

This fear of complications and death sometimes translates to a fear of going to the doctor and addressing concerns about their own health.

> *“I feel scared to discuss with them, I feel they might say something new. Now I am already suffering from a lot, I don’t want to suffer even more.”* (P9, Female, PHQ-9 = 15, Moderately-Severe Depression)

Many participants expressed sentiments of feeling like a burden to their family members:

> *“I definitely feel that I am becoming a trouble to someone. I also feel I should not depend on anyone, neither my husband nor son. I should take care of myself.”* (P12, Female, PHQ-9 = 7, Mild Depression)

For some, this feeling had built over time to suicidal thoughts.

> *“I feel that why should I live, all are facing problems because of me. No one will take care of me except my husband. So I feel I shouldn’t be here, why should everyone suffer because of me?”* (P8, Female, PHQ-9 = 9, Mild Depression)

Participants reported a significant impact to their energy levels and working capacity since their diagnosis with diabetes:

> *“I am not able to work as much as others, I get stressed quickly. I feel fatigued. I have to take tablets on time, and after taking them, I feel really fatigued. If I happen to take the tablets half an hour later because of some chores around the house, I will feel really fatigued. I feel like eating immediately. I am not able to walk like others. People who are my age and don’t have any sugar [diabetes], they can walk well. I do not have as much strength as them.”* (P15, Female, PHQ-9 = 6, Mild Depression)

## Discussion

With the rising prevalence of T2D in younger populations, it is critical to understand their lived experiences and identify barriers to their effective diabetes management and overall well-being. This study concurrently explores the occurrence of depressive symptoms with the lived experiences, facilitators, and barriers to T2D-related self-care activities in young adults under the age of 35 in Mysore district, South India. Overall, participants exhibited high rates of depressive symptoms and suicidal ideation. Key barriers and facilitators to diabetes self-management and overall quality of life include knowledge and awareness about the condition, experiences with and in the healthcare system, presence of depressive symptoms, and physical and external manifestations and impact of the condition on other aspects of their lives.

All study participants described challenging circumstances in their daily lives with the condition, regardless of their PHQ-9 score. Negative feelings towards the condition, feeling like a burden, and worry about the future were sentiments expressed by participants at all levels of depression symptoms. In this study, 14 participants (70%) presented mild to moderately-severe depressive symptoms, with 5 participants (25%) reporting recent suicidal thoughts. Compared to the national prevalence estimate of 3.6% in India, young adults with T2D are at a greater risk of depressive disorders [28]. Descriptively in this sample, depressive symptoms are associated with sex, region, socioeconomic status, duration since T2D diagnosis, family history of T2D, T2D-related complications, adherence to a diabetic diet, and physical activity. This data is consistent with the established relationship between diabetes and depressive symptoms recognized around the world as well as factors shown to be associated with this relationship [7,8,10–14]. All people living with diabetes must manage the constant, daily effects associated with the condition. Chronic illnesses are challenging for everyone living with them, both physically and mentally, and it is important that both are treated concurrently to improve overall health [12].

The interviews revealed several knowledge gaps and misconceptions related to diabetes. Many participants lacked trustworthy sources of information about the condition and were afraid to ask questions due to the stigma associated with it. The belief that diabetes is communicable was particularly prevalent and concerning, leading to experiences of social isolation and discrimination. Researchers in the United States and Ghana have established that lower health literacy coupled with illness perception is associated with lower self-management behaviors in adults living with T2D [28–30]. Medication perception also played a significant role in participants’ experience with T2D. Most participants expressed reluctance to rely on medication for the rest of their lives, influenced by both healthcare professionals and community members. Similar results were found by Miglani et al. in India (2000), where young adults living with diabetes reported fear that people would treat them differently and experienced unsolicited advice and instructions about household remedies from family members and peers [17]. Researchers in Saudi Arabia and Japan have found that medication adherence is dependent on the beliefs toward the medications and potential T2D complications [31,32]. This speaks to the urgency of implementing public health campaigns to dispel myths circulating within local communities and provide accurate information about T2D, including the importance of medication adherence for long-term glycemic control and prevention of future complications. Adults with T2D are more likely to form positive perceptions about T2D with increased health literacy, encouraging self-care practices for glycemic control, eliminating cultural, social, and personal convictions about the condition, and building patient compliance and self-efficacy [33].

The healthcare system plays a pivotal role in T2D management. Participants reported mixed interactions with healthcare providers; while they generally recommended lifestyle modifications, including diet and exercise, individualized care and evidence-based support was often lacking. In some cases, participants experienced blame and scolding by their healthcare providers, which is shown to have detrimental effects on patient motivation and self-efficacy [34,35]. Complex power dynamics between patients and providers exist in both urban and rural Mysore district. A qualitative study conducted in rural Australia found that power dynamics and trust between healthcare providers and patients greatly influence patient experiences and perceived care [36]. Furthermore, rural participants’ preference for private clinics underscores the need for improved accessibility and quality of public healthcare services. Similar findings were established in Vietnam, where researchers found that social factors including word-of-mouth, relationships with healthcare providers, healthcare staff attitudes, and marketing influence the choice of private over public services [37].

Self-care activities are essential components of diabetes management, but participants expressed numerous barriers that they experience. Physical activity was hindered by fatigue, concerns about social judgment, and a lack of time. Healthy diets were limited by financial constraints and family priorities, as female participants expressed difficulties in maintaining separate diets from their family members.

Medication adherence was also influenced by financial constraints, as some participants had to choose between purchasing their medication or investing in something else. Previous research amongst Asian young adults in China and Singapore recognized that adherence to diabetes medication is poorer in younger adults than in older adults [28]. Researchers found similar results in urban slums in South India, where the overall prevalence of self-care activities was low, indicating a need for more awareness and education about diabetes self-care management [38]. South Asian populations with T2D have been shown to experience these barriers to self-care activities, including communication discordance, lack of knowledge and misperceptions about the condition and management, gender-specific roles, lack of family support and cultural appropriateness [39]. Addressing these barriers requires a multifaceted approach, including community support, increased awareness, and access to safe physical activity opportunities, healthy foods, and affordable medications.

The presence or lack of social support played a critical role in the participants’ diabetes management. Supportive spouses and friends provided encouragement and motivation to some participants, emphasizing the importance of involving peers in diabetes care. Social and family support has been shown to improve motivation, self-efficacy, and health outcomes in people living with T2D around the world [35,40,41]. However, competing priorities, particularly related to children’s education, often took precedence over self-care. Previous research has shown that occupation, living condition, comorbidity, current treatment, and history of hospitalization are also predictors of perceived family burden of T2D [42]. It is important that young adults with T2D feel empowered to prioritize their health, without feeling like a burden to their families; increased social support amongst this rising population has the potential to build community and foster solidarity to improve both physical and mental health outcomes.

## Strengths and limitations

A major strength of this study is the collaboration with local researchers in Mysore district, who had existing trust and relationships within the community. All interviews were conducted by one research assistant to maintain internal validity and consistency across the interviews. Qualitative analysis was conducted by three researchers; one participated in the interviews to account for contextual cues and the other two were not present, minimizing bias and allowing for objectivity in the outcomes. The local researchers were consulted closely to ensure adequate representation of the local context in the presented qualitative and quantitative findings.

Thematic saturation was reached with the sample of qualitative interviews, but it was too small for robust quantitative analysis. There is inherent selection bias in who chose to participate in the study, which may have positively skewed the results. Additionally, there is gender asymmetry in the study participants; while we planned to recruit an equal number of male and female T2D patients, more females volunteered for the study. Some results may be biased by participants’ perceptions of health research, but we employed probing to minimize this bias as much as possible. Causal inferences cannot be drawn, given the study’s cross-sectional design. However, the purpose of this study was not for generalizability, but rather an in-depth exploration of lived experiences with T2D in Mysore district.

## Future directions

Future research should consider larger and more diverse samples, from both urban and rural Mysore district. Longitudinal studies could provide insights into how young adults’ experiences and perceptions of diabetes change over time. Additionally, interventions tailored to address the identified barriers to self-care activities and psychosocial well-being should be developed and evaluated for their effectiveness in improving T2D management and overall quality of life in young adult populations.

## Conclusion

The findings of this study highlight the multifaceted challenges faced by young adults living with T2D in Mysore district, South India. Addressing these challenges requires a holistic approach, including improved healthcare access, education and awareness campaigns, mental health support, and interventions to overcome barriers to self-care practices. As T2D becomes more prevalent across the world at younger ages, it is imperative that we understand their lived experiences to best support them. Young adulthood is an excellent opportunity to intervene and promote healthy behaviors before life-changing complications occur. By understanding the unique experiences and perceptions of young adults with T2D, healthcare systems and policymakers can develop more effective strategies to support diabetes management and improve the long-term well-being of those affected.

## Data Availability

The availability of the full data set is not publicly available in a repository due to ethical restrictions. Ethical approval for the study was received from Duke Campus Institutional Review Board (2022-0358) and the Public Health Research Institute of India’s Institution Ethics Review Board (2022-01-29-64). When applying for ethical approval we did not specify that the data would be made publicly available in a repository. As part of the written and verbal consent we assured participants that all data would be confidential and access to the recordings would be restricted to the research team. We did specify that “some of their words” may be used in reporting the findings of the study, which we have done within the manuscript as non-identifiable quotes).

